# Genetic liability for substance use associated with medical comorbidities in electronic health records of African- and European-ancestry individuals

**DOI:** 10.1101/2021.04.19.21255619

**Authors:** Emily E. Hartwell, Alison K. Merikangas, Shefali S. Verma, Marylyn D. Ritchie, Regeneron Genetics Center, Henry R. Kranzler, Rachel L. Kember

## Abstract

Polygenic risk scores (PRS) represent an individual’s summed genetic risk for a trait and can serve as biomarkers for disease. Less is known about the utility of PRS as a means to quantify genetic risk for substance use disorders (SUDs) than for many other traits. Nonetheless, the growth of large, electronic health record-based biobanks makes it possible to evaluate the association of SUD PRS with other traits. We calculated PRS for smoking initiation, alcohol use disorder (AUD), and opioid use disorder (OUD) using summary statistics from the Million Veteran Program sample. We then tested the association of each PRS with its primary phenotype in the Penn Medicine BioBank (PMBB) using all available genotyped participants of African or European ancestry (AFR and EUR, respectively) (N=18,612). Finally, we conducted phenome-wide association analyses (PheWAS) separately by ancestry and sex to test for associations across disease categories. Tobacco use disorder was the most common SUD in the PMBB, followed by AUD and OUD, consistent with the population prevalence of these disorders. All PRS were associated with their primary phenotype in both ancestry groups. PheWAS results yielded cross-trait associations across multiple domains, including psychiatric disorders and medical conditions. SUD PRS were associated with their primary phenotypes, however they are not yet predictive enough to be useful diagnostically. The cross-trait associations of the SUD PRS are indicative of a broader genetic liability. Future work should extend findings to additional population groups and for other substances of abuse.

## Introduction

Substance use disorders (SUDs) are prevalent and costly to society. In 2019, 7.8% of U.S. adults had a current (past-year; PY) SUD.^1^ These disorders are associated with a host of negative outcomes, including poorer quality of life and increased mortality risk.^2-4^ The most common SUDs are tobacco use disorder (TUD; 26.9% PY) and alcohol use disorder (AUD; 13.9% PY).^5,6^ Although opioid use disorder (OUD) is less prevalent (0.6% PY) than either TUD or AUD, in 2019, 10.1 million individuals endorsed PY opioid misuse,^1^ which is of particular public health concern due to the associated high risk of fatal opioid overdose.^7^

Evidence from twin studies support a genetic component to the etiology of SUDs, and genome-wide association studies (GWAS) have identified dozens of variants contributing to SUD risk.^8-10^ Nonetheless, because SUDs are polygenic traits, no single variant accounts for more than a small portion of the variance in risk for developing an SUD. Polygenic risk scores (PRS) calculated from the weighted effect size of the GWAS-derived variants associated with a trait provide an aggregate measure of common genetic risk.^11^ PRS have been used to estimate the risk of medical diseases,^12^ health outcomes,^13^ and psychiatric disorders.^14^

Additionally, PRS have been used to investigate the genetic overlap between multiple phenotypes, which can help to evaluate overlapping and unique features of SUDs, that share symptomatology and, to some extent, underlying genetic architecture.^15^ Consistent with these features, a study of 11 different psychiatric and SUDs GWAS yielded strong genetic correlations between alcohol and tobacco dependence and other psychiatric disorders.^16^

Electronic health records (EHRs) are integral to the investigation of co-occurring disorders, as they provide a wealth of longitudinal phenotypic information that is often richer than that collected in clinical trials or cross-sectional studies. This longitudinal perspective, which provide repeated measures of a trait over long periods of time, can thereby increase confidence in the accuracy of diagnoses. Biobanks that provide genetic data linked with EHRs have been the basis for the development of phenome-wide association studies (PheWAS).^17^

PheWAS can use genetic data, such as a genetic variant or a PRS, to query phenotypic information (e.g., medical or psychiatric diagnoses, environmental factors) to identify associations, thus potentially illuminating shared genetic etiology or yet undiscovered phenotypic comorbidity. Although the use of GWAS and PheWAS has led to the identification of thousands of disease-associated variants, the vast majority of this literature comes from studies of individuals of European ancestry, underscoring the substantial presence of health disparities in genetic research. Women have also historically been underrepresented in medical research. The Penn Medicine BioBank (PMBB), like a number of other biobanks, is well suited to address these gaps in the genetics literature given the heterogeneity of its participants, who are drawn from a large urban center with a diverse population.

Using available summary statistics, we calculated PRS for three common substance-related traits; smoking initiation, AUD, and OUD, in both African-ancestry (AFR) and European-ancestry (EUR) individuals and examined their performance and their phenotypic associations in the PMBB. We also performed PheWAS to identify cross-trait associations and phenotypic overlap in genetic liability for SUDs, and a secondary PheWAS where the primary phenotype was covaried to determine whether identified associations persist when controlling for the index phenotype. Lastly, we conducted PheWAS separately in men and women to test whether the associations differed by sex.

## Methods

### Participants

Patients in the Penn Medicine BioBank (PMBB) were ascertained at the time of a medical appointment in the University of Pennsylvania Health System. At enrollment, participants gave informed consent and provided either a blood or tissue sample and permission to access their EHR information. As of July 2020, there were 63,177 individuals in PMBB, 21,263 of whom had genome-wide genotyping.

### Genotyping and Quality Control

DNA was extracted from blood and samples were genotyped in four batches: 1) N=10,867 using the Illumina InfiniumOmniExpress-24v1-2_A1 (OMNI) at the Regeneron Genetics Center (RGC); 2) N=5,676, using the Global Screening Array (GSA) V1 at the Center for Applied Genomics (CAG) at the Children’s Hospital of Philadelphia; 3) N=2,972, using the GSA V2 chip at CAG; and 4) N=16,940 using the GSA V2 chip, at RGC. Some samples were genotyped multiple times. To impute unique samples, they were combined by genotyping chip into two batches (3 GSA chips into a GSA batch and 1 OMNI chip into an OMNI batch), with preference for unique samples in the following order: GSA V2, GSA V1, OMNI. Each batch underwent quality control separately prior to imputation. Samples were removed if the genotyping call rate was <90% or if genotyped sex did not match reported gender, leaving 20,079 unique GSA samples and 1,111 unique OMNI samples. SNPs were removed if the call rate was <95%, leaving N=622,717 SNPs in GSA and N=640,714 SNPs in OMNI.

### Phasing and Imputation

Prior to imputation, SNPs were matched to the appropriate strand and those that were palindromic, not matched to the reference panel, with allele frequency difference >0.2, or with differing alleles were removed. Genotypes were phased (using EAGLE) and imputed to the TOPMed Reference Panel (Freeze 5) on the TOPMed Imputation server.^18^ In the GSA batch, 151,143,913 SNPs were imputed with R^2^>0.3. In the OMNI batch, 46,386,520 SNPs were imputed with R^2^>0.3.

Imputed genotypes underwent quality control (R^2^>0.3, marker call rate>95%, sample call rate>90%, MAF>0.5%) using PLINK v1.90. Related individuals were identified using a graph-based algorithm after applying a Pi-HAT threshold of 0.25, and the sample most closely related to multiple other samples was removed (n=738). Following removal of related individuals, principal component (PC) analysis was conducted using the Eigensoft smartpca module.^19^ We performed quantitative discriminant analysis to determine genetically informed ancestry, using 1000 Genomes, phase 3^20^ as a training set and PMBB as a testing set.

### Polygenic Risk Scores

We retained SNPs present in the HapMap reference panel (n=1,120,629) and merged the two batches into a single dataset. We extracted AFR (n=8,276) and EUR (n=10,473) individuals and used PLINK v1.90 to calculate ancestry-specific PCs to use as covariates. Summary statistics for smoking initiation (current vs. never smokers, Contrast I),^10^ AUD (AFR^8^; EUR^21^), and OUD^9^ were obtained from the Million Veteran Program (MVP) (Supplemental Table S1). The MVP is one of the largest available sources for GWAS summary statistics for these phenotypes and, importantly, is the largest available sample of AFR individuals. Ancestry specific PRS were calculated using PRS-CS,^22^ with AFR GWAS summary statistics and the 1000G AFR LD reference panel used in the PMBB AFR individuals, and the EUR GWAS summary statistics and the 1000G EUR LD reference panel used in the PMBB EUR individuals. Phi for all traits was fixed to 1e-2, with default thresholds used for everything else.

### Phenotypes

International Classification of Diseases (ICD)-9 and ICD-10 data for 63,199 individuals were extracted from the EHR and used to assign both primary phenotypes and phecodes. To increase the accuracy of diagnostic data, encounter type was filtered to include only records representing physician encounters, as previously described.^14^ This left 11,966,749 encounters (5,731,365 ICD-9; 6,235,384 ICD-10) for 63,177 individuals. ICD codes for TUD, AUD, and OUD were selected based on clinician expertise (Supplemental Table S2). We classified <u>primary phenotypes</u> in two ways: a less stringent definition (requiring the presence of ≥1 or more ICD codes for the phenotype), and a stringent definition (requiring the presence of ≥1 ICD codes as inpatient, or ≥2 as outpatient). To create a dataset for PheWAS analysis, we aggregated ICD-9 codes to <u>phecodes</u> using the phecode ICD-9 map 1.2 and ICD-10 codes using the phecode ICD-10-CM map 1.2 (beta). Individuals were considered cases for the phecode if it was assigned on at least 2 unique dates, controls if they have no instance of the phecode, and ‘other/missing’ if they had one instance or a related phecode.^23^ The final dataset included 18,612 individuals (8,235 AAs, 10,377 EAs) with complete genotype, phenotype, and covariate data.

### Statistical Analysis

Individuals with a given SUD and those without that SUD were compared on demographics and comorbid medical conditions via chi-square for categorical data and t-tests for continuous data. PRS were standardized with mean=0 and standard deviation (SD)=1. Logistic regression was used to test for the association of each psychiatric risk score with the primary phenotype and to estimate the odds ratio (OR) for cases by comparing the top quintile of polygenic risk to the remaining quintiles of risk. We used TUD as the primary phenotype for smoking initiation, as it was the most similar one available in the EHR. The PheWAS analysis was performed in R using logistic regression models in which each PRS was the independent variable; phecodes were the dependent variables; and age, sex, and the first 10 PCs were covariates. We performed a second PheWAS that adjusted for the primary phenotype by including it as a covariate in the regression model and a third PheWAS that tested for sex differences. Phecodes with >100 cases (n=583 for AFR; n=477 for EUR) were tested (see Supplement 2). As a different number of Phecodes were tested in each ancestry, Bonferroni-corrected phenome-wide significance thresholds to account for multiple testing were p<8.59x 10^−5^ for AFR and p<1.05×10^−4^ for EUR.

## Results

### Phenotypes in PMBB

Participants’ demographics and information on their comorbid conditions are presented in Table 1 for the genotyped sample (n=18,612) by SUD and ancestry. The sample was 53% male, with a mean age of 65 (SD=16.7) years, and a mean of 61 (SD=9.9) encounters in the EHR. As anticipated due to the initial PMBB recruitment strategy, there were high rates of comorbid circulatory system (82%), endocrine/metabolic (73.5%) and respiratory system (51%) diagnoses. Notably, individuals with SUDs had significantly higher rates of comorbid conditions, irrespective of SUD or ancestry.

**Table 1.**
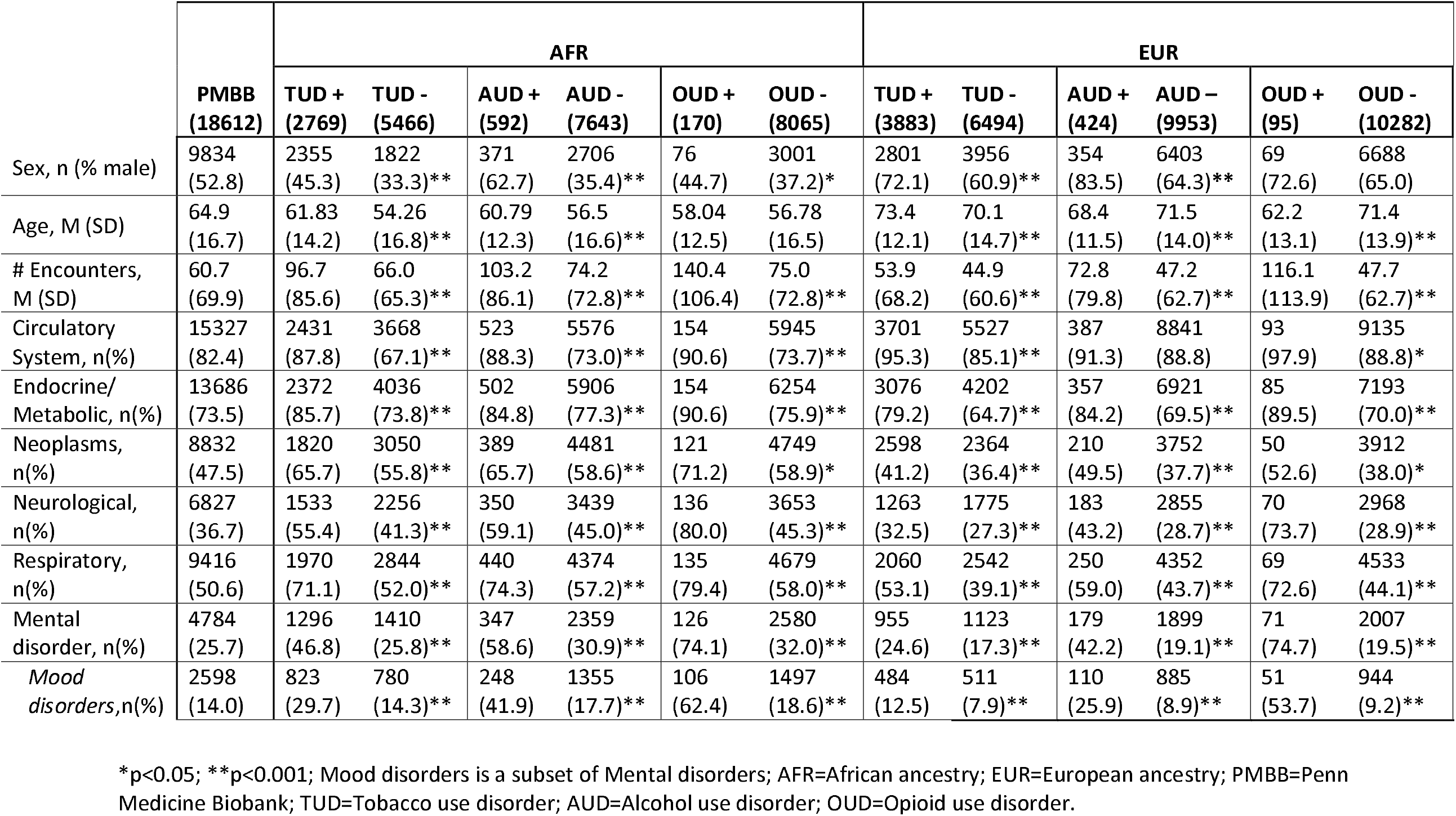
Demographic information and prevalence of key PheCode comorbidities

### Primary associations of SUD PRS

PRS were calculated for smoking initiation, AUD, and OUD from summary statistics available from GWAS conducted in the Million Veteran Program (Supplemental Table S1). We tested the association of the three PRS with their respective primary phenotypes, using both less stringent and stringent case definitions (Table 2). Using the less stringent case definition, PRS_Smoking_ was strongly positively associated with TUD in both the AFR and EUR samples (AFR cases = 2,769, OR = 1.12, *p* = 1.4 × 10^−8^; EUR cases = 3,883, OR = 1.25, *p* = 3.6 × 10^−22^). Likewise, the PRS_AUD_ was positively associated with AUD in both the AFR and EUR samples (AFR cases = 592, OR = 1.09, *p* = 0.04; EUR cases = 424, OR = 1.19, *p* = 7.1 × 10^−4^) as was PRS_OUD_ (AFR cases = 170, OR = 1.24, *p* = 0.01; EUR cases = 95, OR = 1.28, *p* = 0.02). Similar associations were obtained using the stringent phenotype definition, although PRS_OUD_ was no longer significant in the EUR sample (Table 2).

**Table 2.**
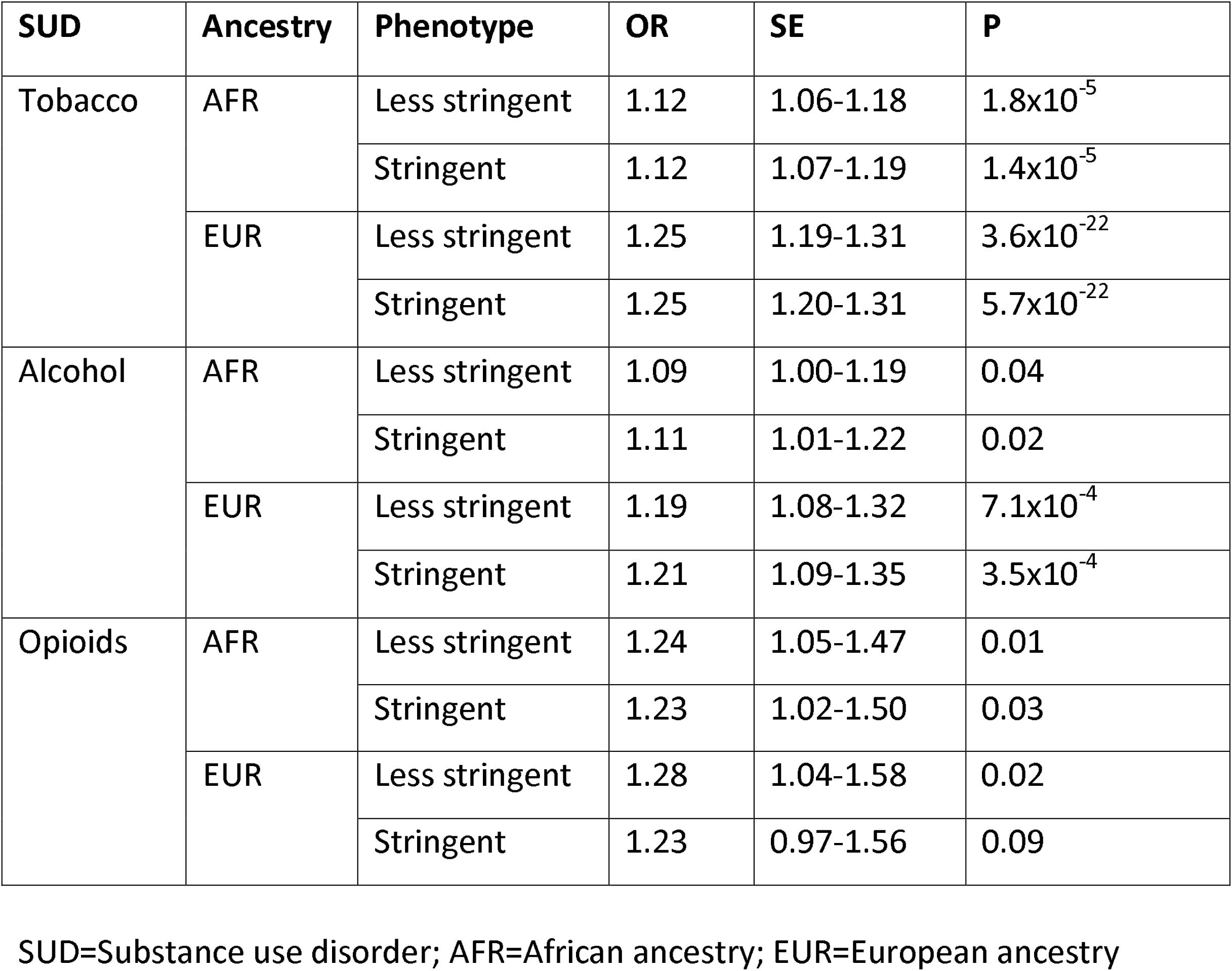
Polygenic risk scores results by substance use using the PRS-CS method

| **SUD** | **Ancestry** | **Phenotype** | **OR** | **SE** | **P** |
| --- | --- | --- | --- | --- | --- |
| Tobacco | AFR | Less stringent | 1.12 | 1.06-1.18 | $1.8 \times 10^{-5}$ |
| | | Stringent | 1.12 | 1.07-1.19 | $1.4 \times 10^{-5}$ |
| | EUR | Less stringent | 1.25 | 1.19-1.31 | $3.6 \times 10^{-22}$ |
| | | Stringent | 1.25 | 1.20-1.31 | $5.7 \times 10^{-22}$ |
| Alcohol | AFR | Less stringent | 1.09 | 1.00-1.19 | 0.04 |
|  |  | Stringent | 1.11 | 1.01-1.22 | 0.02 |
| | EUR | Less stringent | 1.19 | 1.08-1.32 | $7.1 \times 10^{-4}$ |
| | | Stringent | 1.21 | 1.09-1.35 | $3.5 \times 10^{-4}$ |
| Opioids | AFR | Less stringent | 1.24 | 1.05-1.47 | 0.01 |
|  |  | Stringent | 1.23 | 1.02-1.50 | 0.03 |
|  | EUR | Less stringent | 1.28 | 1.04-1.58 | 0.02 |
|  |  | Stringent | 1.23 | 0.97-1.56 | 0.09 |

SUD=Substance use disorder; AFR=African ancestry; EUR=European ancestry

For each phenotype, we determined the case prevalence by PRS quintile. PRS_Smoking_ showed an absolute risk of 36.7% for AFR and 44.5% for EUR in the top quintile (Figure 1), with corresponding odds ratio (OR) of 1.20 (*p* = 0.004) for AFR and 1.45 (*p* = 3.6×10^−13^) (Supplemental Table S3). The top quintile for PRS_AUD_ showed an absolute risk of 7.8% for AFR (OR = 1.13, *p* = 0.25) and 6.1% for EUR (OR = 1.59, *p* = 5.2×10^−5^) (Figure 1; Supplemental Table S3). For PRS_OUD_, the absolute risk in the top quintile was 2.5% for AFR (OR = 1.28, *p* = 0.21) and 1.3% for EUR (OR = 1.64, *p* = 0.03) (Figure 1; Supplemental Table S3).

**Figure 1.**
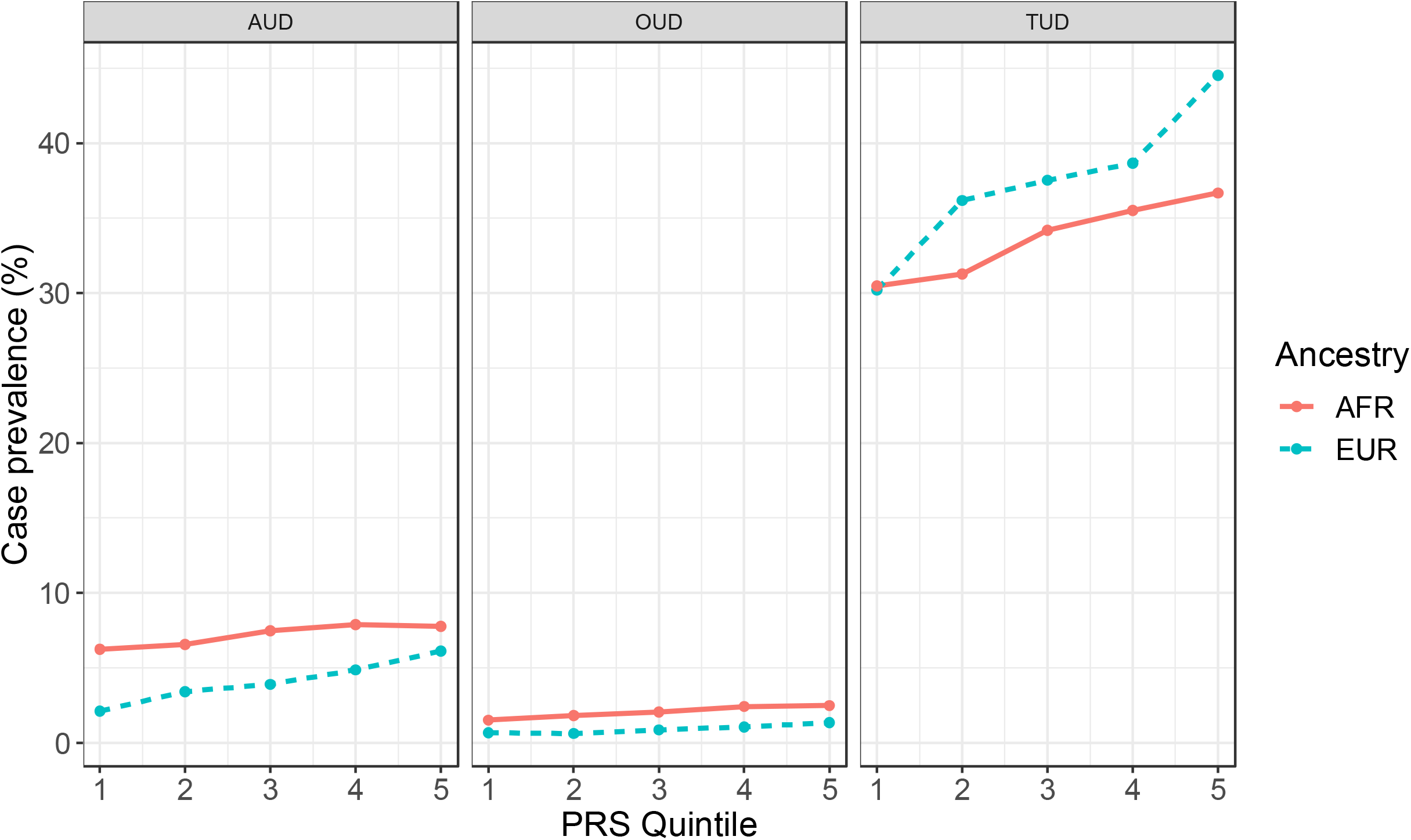
Case prevalence (%) by PRS quintile for AUD, OUD, and TUD using the less stringent definition

### Phenome-wide analysis of SUD PRS

#### Tobacco

There was a significant association of the AFR PRS_Smoking_ with TUD (OR = 1.19, *p* = 4.95×10^−9^) and strong, though after Bonferroni correction not statistically significant, associations with emphysema (OR = 1.34, *p* = 1.3×10^−4^), chronic airway obstruction (OR = 1.17, *p* = 2.8×10^−4^), alcohol-related disorders (OR = 1.26, *p* = 2.9×10^−4^) and alcoholism (OR =1.29, *p* = 3.0×10^−4^) (Figure 2A). Covarying for TUD, the AFR PRS_Smoking_ showed no statistically significant associations (Figure 2B). The analysis by sex (Supplemental Figure S1) showed there were strong associations of the PRS_Smoking_ with TUD in both men (OR = 1.23, *p*= 2.46×10^−5^) and women (OR = 1.18, *p* = 3.27×10^−5^).

**Figure 2.**
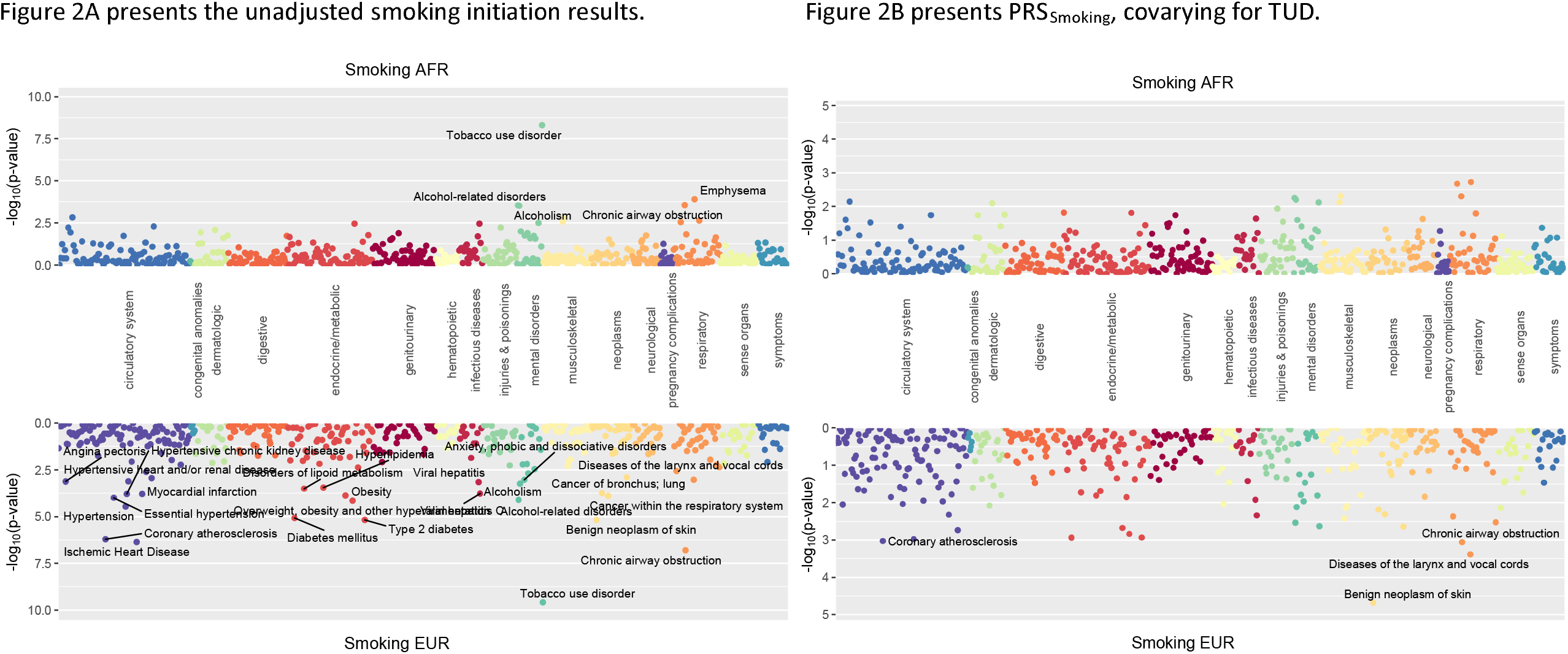
Phenome-wide association of polygenic risk scores for smoking initiation.

The EUR PRS_Smoking_ was significantly associated with 12 phenotypes, including increased risk of TUD (OR = 1.25, *p* = 2.61×10^−10^), chronic airway obstruction (OR = 1.21, *p* = 1.57×10^−7^), ischemic heart disease (OR = 1.14, *p* = 4.43×10^−7^), coronary atherosclerosis (OR = 1.14, *p* = 6.11×10^−7^) and Type 2 diabetes (OR = 1.14, *p* = 6.53×10^−6^) (Figure 2A). Covarying TUD, the only association that remained significant was a protective effect against benign neoplasm of skin (OR = 0.88, *p* = 2.09×10^−5^) (Figure 2B). Analysis by sex (Supplemental Figure S1) showed that, in men, PRS_Smoking_ was significantly associated with 10 phenotypes, including TUD (OR = 1.24, *p* = 1.24×10^−7^), chronic airway obstruction (OR = 1.22, *p* = 8.67×10^−6^), several circulatory system diseases (e.g., hypertension, OR = 1.16, *p* = 1.57×10^−5^), and Type 2 diabetes (OR= 1.16, *p* = 1.87×10^−5^). In women, PRS_Smoking_ remained strongly associated with TUD (OR = 1.28, *p* = 7.22×10^−5^), with smaller associations with thyroid disorders (OR = 0.62, *p* = 4.8×10^−4^), benign neoplasm of skin (OR = 0.86, *p* = 5.3×10^−4^).

#### Alcohol

The AFR PRS_AUD_ was not significantly associated with any phenotype (Figure 3A), which was not altered by adding AUD as a covariate (Figure 3B) or examining PRS_AUD_ by sex (Supplemental Figure S2).

**Figure 3.**
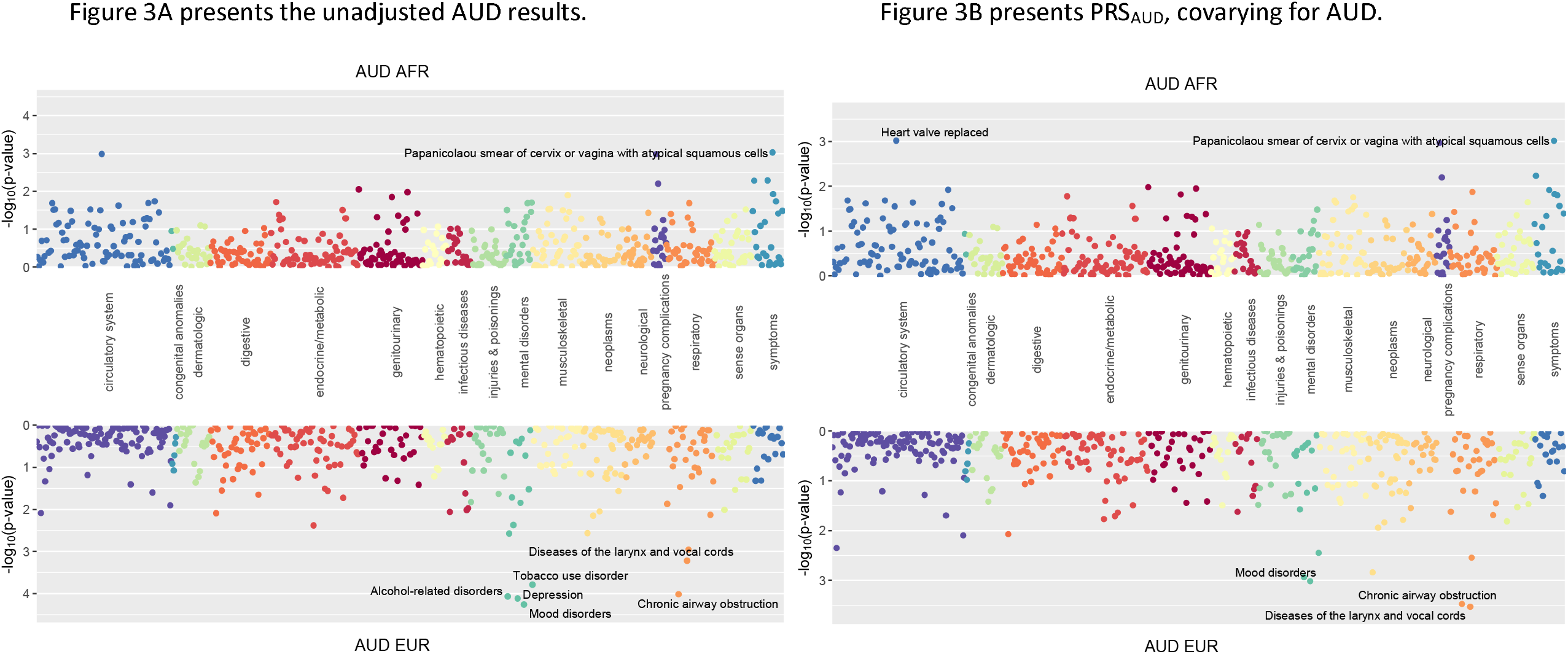
Phenome-wide association of polygenic risk scores for alcohol use disorder (AUD).

The EUR PRS_AUD_ showed significant positive associations with mood disorders (OR = 1.16, *p* = 5.49×10^−5^), depression (OR = 1.16, *p* = 7.65×10^−5^), alcohol-related disorders (OR = 1.35, *p* = 8.49×10^−5^), and chronic airway obstruction (OR = 1.15, *p* = 9.63×10^−5^) (Figure 3A). Further, it was nominally positively associated with TUD (OR = 1.14, *p* = 1.62×10^−4^). Covarying AUD demonstrated a nominally significant relationships with protective effect for disease of the larynx and vocal chords (OR = 0.78, *p* = 2.97×10^−4^), chronic airway obstruction (OR = 1.13, *p* = 3.36×10^−4^) and mood disorders (OR = 1.13, *p* = 9.62×10^−4^) (Figure 3B). In men, there were strong positive associations with mood disorders (OR = 1.25, *p* = 1.12×10^−5^), depression (OR = 1.25, *p* = 1.54×10^−5^), and alcohol-related disorders (OR = 1.43, *p* = 2.06×10^−5^), but no strong associations in women (Supplemental Figure S2).

### Opioids

The AFR PRS_OUD_ was not significantly associated with any phenotype (Figure 4A), though there were nominal, positive associations with fracture of ankle and foot (OR = 1.31, *p* = 1.56×10^−3^) and acute pulmonary heart disease (OR = 1.22, *p* = 3.48×10^−3^). A similar pattern of findings was obtained when OUD was included as a covariate (Figure 4B). Analysis by sex showed no significant associations (Supplemental Figures S3A and S3B).

**Figure 4.**
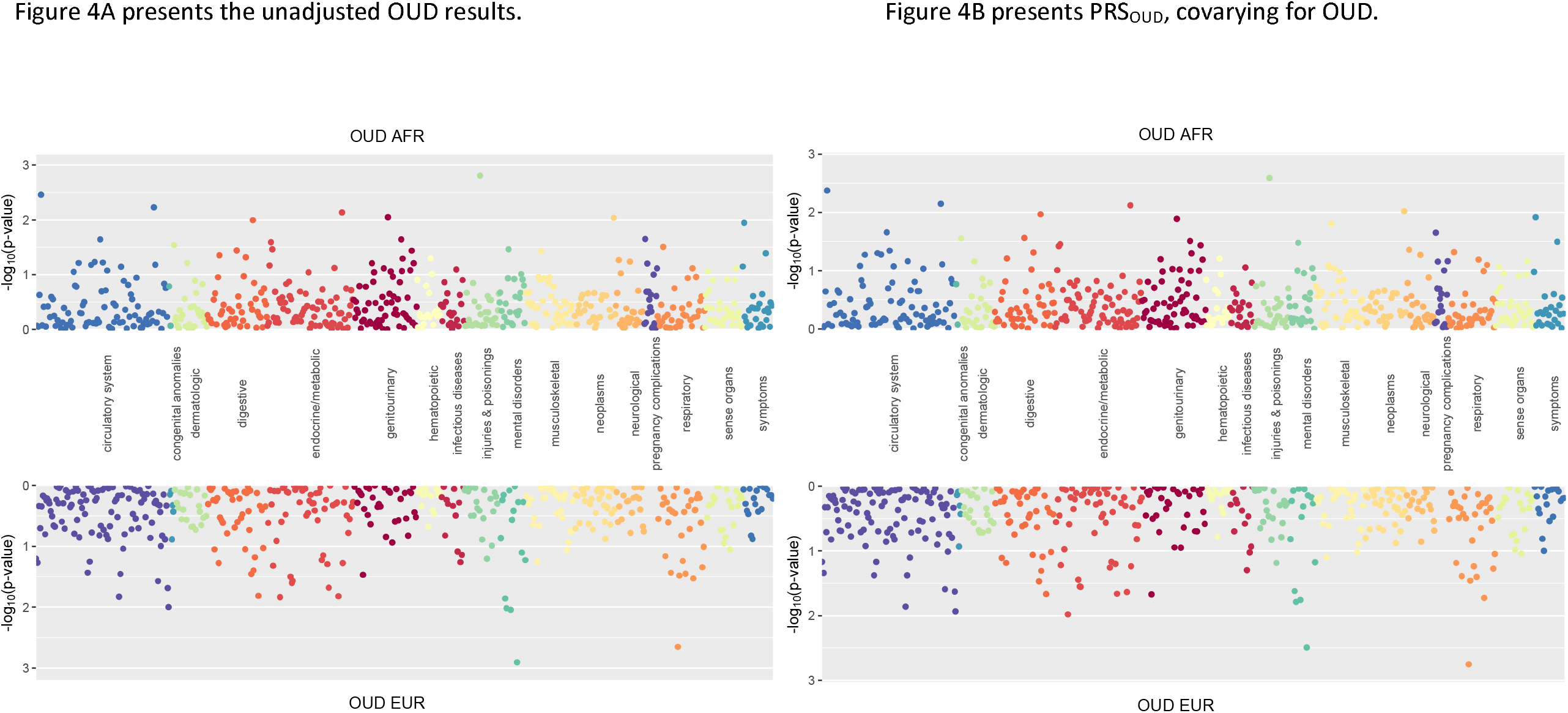
Phenome-wide association of polygenic risk scores for opioid use disorder (OUD).

The EUR PRS_OUD_ was not significantly associated with any phenotype, though there was a nominally positive association with mood disorders (OR = 1.12, *p* = 1.2×10^−3^) and diseases of the larynx and vocal chords (OR = 0.82, *p* = 2.2×10^−3^) (Figure 4A). Covarying OUD yielded similar results (Figure 4B). When examining PRS by sex, men showed significant positive associations with mood disorders (OR = 1.22, *p* = 3.89×10^−5^) and depression (OR = 1.19, *p* = 3.44×10^−4^) (Supplemental Figure S3A), while there were no significant associations in women (Supplemental Figure S3B).

## Discussion

In this study of 18,612 AFR and EUR individuals from the PMBB, all PRS were associated with their respective diagnoses in both ancestral populations, with the most robust findings being for PRS_Smoking_. The PRS_OUD_ performed less well than the other SUD PRS. The PheWAS results yielded multiple cross-trait associations which have important implications for our understanding of potential shared genetic etiology for comorbid conditions, such as mood and alcohol use disorders. These findings provide new insights into common genetic factors underlying SUD in both AFR and EUR ancestry samples. Given the increasing attention to the lack of diversity in genetic studies,^24^ these findings begin to address the gap in our understanding of the genetic risk factor underlying SUD in African Americans.

The strongest associations were found for PRS_Smoking_, followed by PRS_AUD_ and PRS_OUD_. The difference in the strength of association between PRS and diagnosis is likely a reflection of both the originating GWAS sample size and the prevalence and accuracy of diagnosis in the PMBB dataset. As in previous studies,^25^ the variance explained by PRS for all SUDs was small (e.g., a 13% difference in case prevalence between top and bottom quintile for PRS_Smoking_ in EUR individuals). Thus, while indicative of the risk for the respective SUDs, PRS for the substance-related phenotypes of smoking initiation, AUD, and OUD are currently not adequately predictive to identify cases in an unselected sample.

As expected, in addition to its association with TUD in both ancestral samples, PRS_Smoking_ was associated with alcohol-related disorders and other diseases commonly found in smokers, such as coronary and respiratory disorders. These findings align with both the strong phenotypic relationship between smoking and alcohol use and the known genetic overlap between these traits.^5,10,26^ Interestingly, in both the AFR and EUR samples, these associations were no longer significant after covarying for the primary TUD diagnosis, suggesting that the association between PRS and these phenotypes may be mediated by the TUD diagnosis.

Mirroring known phenotypic comorbidity between AUD and mood disorders,^5^ the EUR sample PRS_AUD_ was associated with mood disorders and depression. This confirms research in other samples that shows that the PRS for MDD is predictive of alcohol dependence.^27^ The association between the PRS_AUD_ and chronic airway obstruction is likely a secondary consequence of smoking, which is also nominally positively associated. These findings replicate the previously observed common genetic factors underlying TUD, alcohol-related disorders, mood disorders, and chronic airway obstruction. However, prior PheWASs of alcohol-related PRS in European samples^8,21^ yielded more significant associations than found here, likely due to their larger sample sizes and increased statistical power.

Likewise, lower statistical power likely explains the lack of significant associations with the PRS_OUD_ PheWAS compared to AUD and TUD in both ancestral samples. In addition, OUD may be under-represented in EHRs in academic medical centers.

There were noteworthy sex differences in our findings, many of which align with established phenotypic patterns regarding SUDs and comorbid conditions. In both men and women, PRS_Smoking_ was significantly associated with TUD. However, there were sex differences in somatic and psychiatric comorbidity; PRS_Smoking_ was not associated with any other mental health conditions in men, whereas it was associated with anxiety disorders in women. This likely reflects the finding that women are more likely to smoke to regulate their mood and mitigate stress^28^ as well as suffer more stress and anxiety from nicotine withdrawal than men.^29^ In contrast, PRS_Smoking_ was associated with circulatory system diseases and Type 2 diabetes in men but not women. This could be because men smoke more cigarettes per day, inhale more deeply, and smoke cigarettes with higher nicotine content in men than in women.^30^

There were also sex differences in the associations with PRS_AUD_, which were associated with mood disorders in EUR men, but not in either EUR or AFR women or AFR men. Given that the comorbidity of mood disorders and SUDs are typically more common in women than men,^31,32^ these findings were not expected. The lack of association of the PRS_AUD_ in women could be attributable to the preponderance of males in the MVP GWAS sample that was used to calculate PRS. Data are mixed regarding whether there is a different genetic liability for alcohol-related outcomes between men and women.^33,34^

The sex-specific association between PRS_OUD_ and mood disorders in EUR men is consistent with findings that mood disorders and OUD are commonly comorbid^32^ and significantly genetically correlated.^9^ As with alcohol and tobacco use, there are sex-specific patterns of opioid use, with younger women using smaller amounts of opioids for shorter periods,^35^ which could either reflect either social or genetic factors.

Limitations of this study include the low prevalence of SUD diagnoses in the PMBB, particularly OUD, which limited statistical power to detect associations. The higher age composition of the PMBB than the general population, associated with a lower prevalence of SUDs, as is typically seen in older adults, likely contributed to the comparatively low prevalence of AUD and OUD. Rates of cardiovascular disease are high in the PMBB because of a concerted effort to recruit from cardiovascular clinics. Therefore, the findings reported here may not generalize to other clinical populations. Finally, the reliance on EHR diagnoses, rather than those obtained through self-report or diagnostic interviews, may have led to misclassification of disorders and the omission of those that occurred outside of Penn Medicine.

The primary strength of this study is the diversity of the PMBB sample, which enabled us to study the large and growing sample of AFR-ancestry individuals in the PMBB. The availability of summary statistics from independent GWAS of AFR-ancestry individuals enabled us to conduct analyses in this understudied population. Another strength of the study is the wealth of phenotypic information available in the Penn Medicine EHR, making it possible to conduct PheWAS. As further GWAS are conducted in diverse ancestral groups, such as those possible in MVP where there are increasing in samples of Latinos and East Asias, and summary statistics become available, we will extend this work to other populations.

This study adds to the growing body of literature demonstrating the genetic liability for SUDs and cross-trait associations with medical and psychiatric comorbidity in AFR- and EUR-ancestry individuals. Identifying differential risk profiles in multiple population groupsand in both sexes may help to interpret the heterogeneity that has challenged our understanding of the genetic architecture of SUD.

## Supporting information

Regeneron Author Listing

Supplement 2 (PheWAS results)

## Data Availability

GWAS summary statistics were contributed by the Million Veteran Program and downloaded from the dbGAP (https://www.ncbi.nlm.nih.gov/projects/gap/cgi-bin/study.cgi?study_id=phs001672.v3.p1). The data, analytic methods, and study materials may be made available to other researchers for purposes of reproducing the results or replicating the procedure upon reasonable request to the corresponding author and with the appropriate ethical approval and data sharing agreements.

## Funding and Acknowledgements

EEH and HRK are supported by the Crescenz VAMC Mental Illness Research, Education and Clinical Center. RLK is supported in part by NIAAA (K01AA028292) and the Million Veteran Program, Office of Research and Development, Veterans Health Administration awards (I01 CX001734 and I01 BX003341). RLK and AKM are supported in part by the TAPITMAT in Translational Medicine and Therapeutics through NCATS (UL1TR001878).

The content is solely the responsibility of the authors and does not necessarily represent the official views of the National Institutes of Health. This publication does not represent the views of the Department of Veterans Affairs or the U.S. government.

We acknowledge and thank the participants of the Penn Medicine Biobank and the Million Veteran Program.

All Regeneron Genetics Center collaborating authors report being employees of Regeneron Pharmaceuticals.

---

**Supplemental Table S1.**
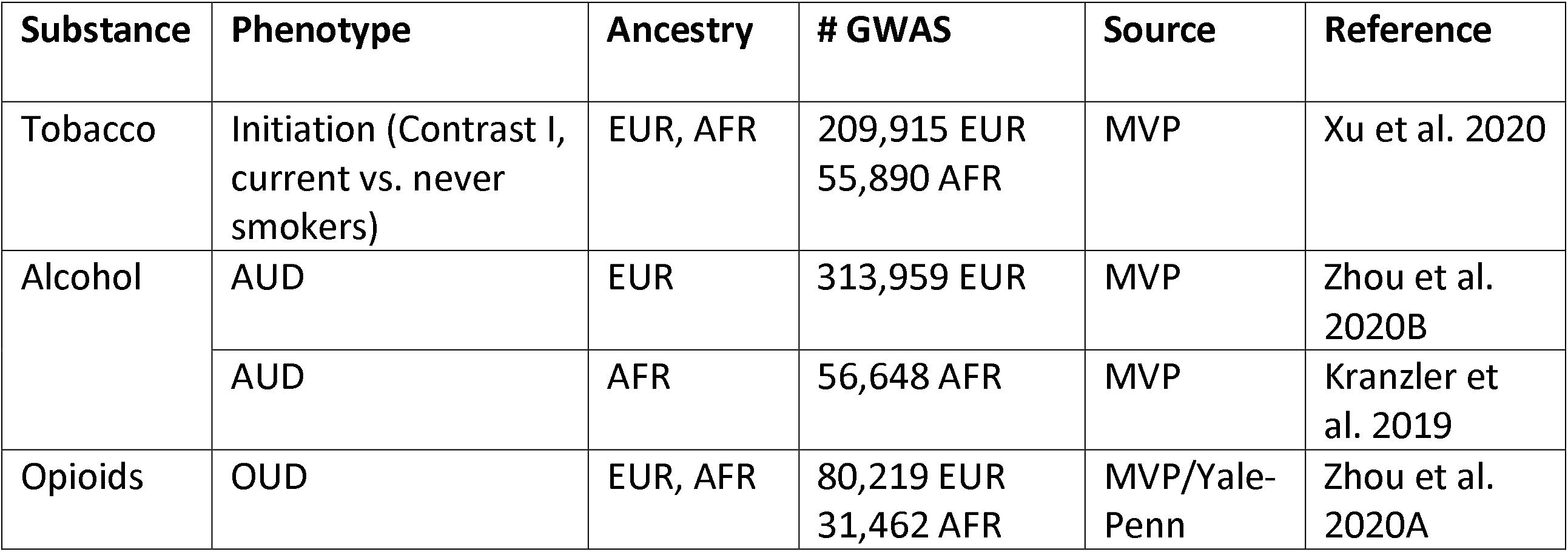
Summary statistic development

**Supplemental Table S2.** ICD codes used to identify cases

| ICD9 | ICD10 | Name |
| --- | --- | --- |
| 305 | F10.1 | Alcohol abuse |
| 303 | F10.2 | Alcohol Dependence |
| 291.81 | F10.3 | Alcohol use, withdrawal |
| -- | F10.9 | Alcohol use, unspecified |
| 571.0, 571.1, 571.2,<br>572.3 | K70-K70.4, K70.9 | Alcoholic liver diseases |
| 535.3 | K29.2 | Alcohol gastritis |
| -- | K86.0 | Alcohol-induced chronic pancreatitis |
| 425.5 | I42.6 | Alcohol cardiomyopathy |
| -- | G31.2 | Degeneration of nervous system due to alcohol |
| -- | G72.1 | Alcohol myopathy |
| V11.2 | -- | Personal history of alcoholism |
| -- | F17.2 | Nicotine Dependence |
| -- | Z72.0 | Tobacco use |
| -- | Z57.31 | Occupational exposure to environmental smoke |
| -- | Z77.22 | Exposure to environmental tobacco smoke |
| -- | Z87.8 | History of nicotine dependence |
| 305.1 | -- | Tobacco Use Disorder |
| 304.0, 304.7 | F11.2 | Opioid Dependence |
| 305.5 | F11.1 | Opioid abuse |

**Supplemental Table S3.** Odds ratios (OR) for top quintile versus the rest for each substance using the less stringent definition of the phenotypes.

| Substance | Ancestry | OR | SE | P |
| --- | --- | --- | --- | --- |
| Tobacco | AFR | 1.20 | 1.06-1.36 | 0.004 |
| | EUR | 1.45 | 1.31-1.61 | $3.6 \times 10^{-13}$ |
| AUD | AFR | 1.13 | 0.92-1.39 | 0.25 |
| | EUR | 1.59 | 1.27-1.99 | $5.2 \times 10^{-5}$ |
| OUD | AFR | 1.28 | 0.87-1.88 | 0.21 |
|  | EUR | 1.64 | 1.04-2.60 | 0.03 |

AFR=African ancestry; EUR=European ancestry; AUD=alcohol use disorder; OUD=opioid use disorder

**Supplemental Figure S1.**
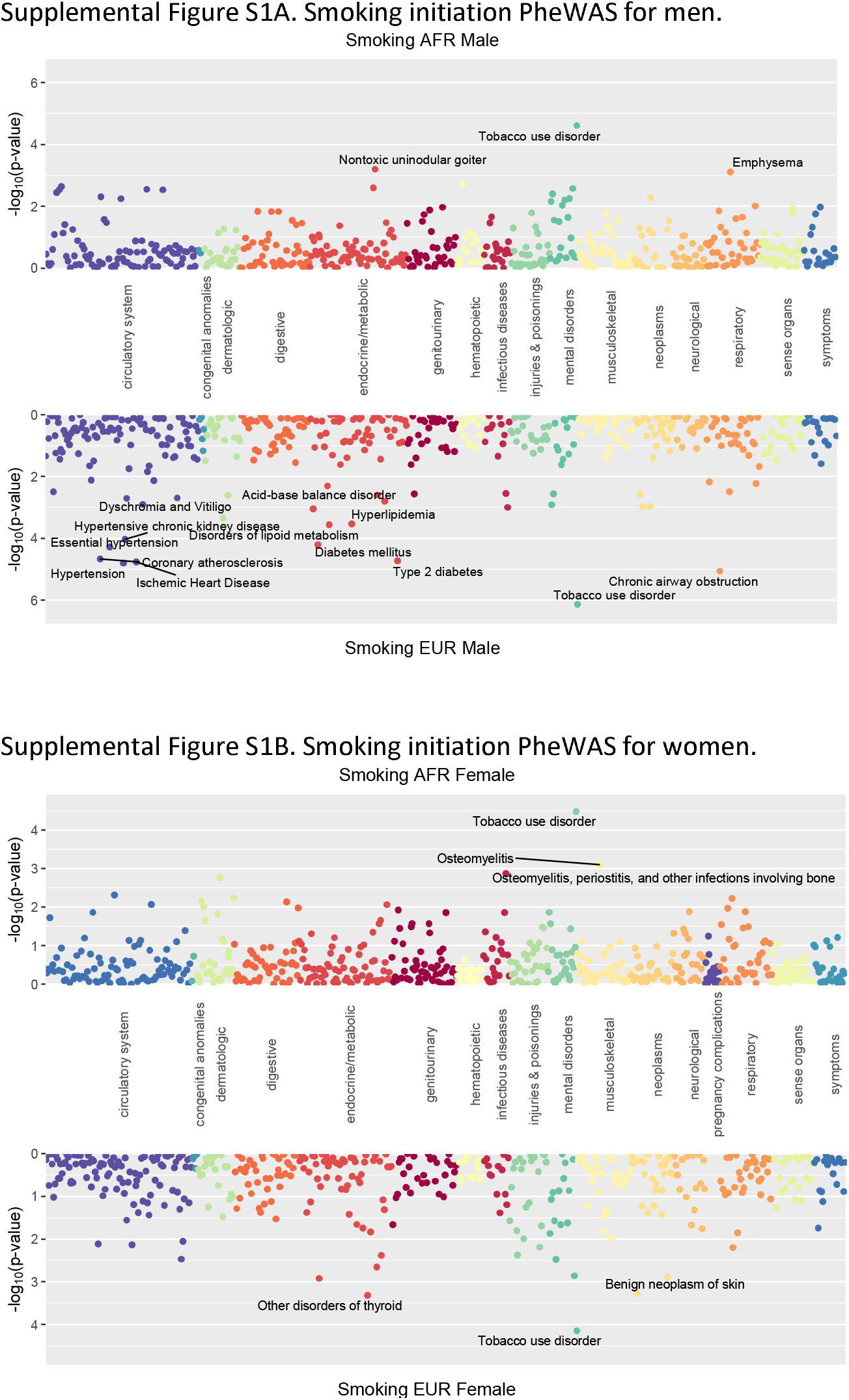
Phenome-wide association of polygenic risk scores for smoking initiation by sex for each ancestry group.

**Supplemental Figure S2.**
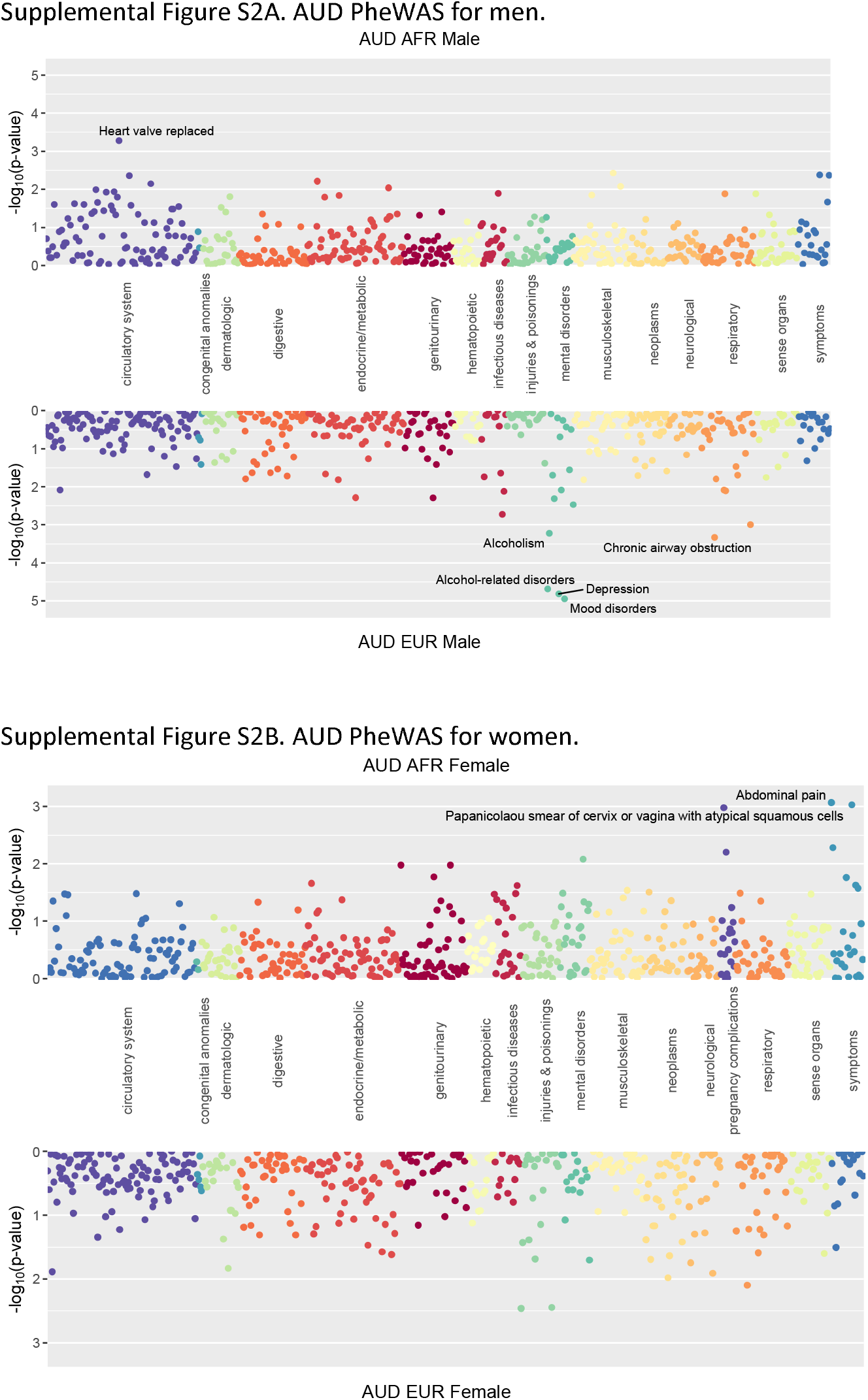
Phenome-wide association of polygenic risk scores for alcohol use disorder (AUD) by sex for each ancestry group.

**Supplemental Figure S3.**
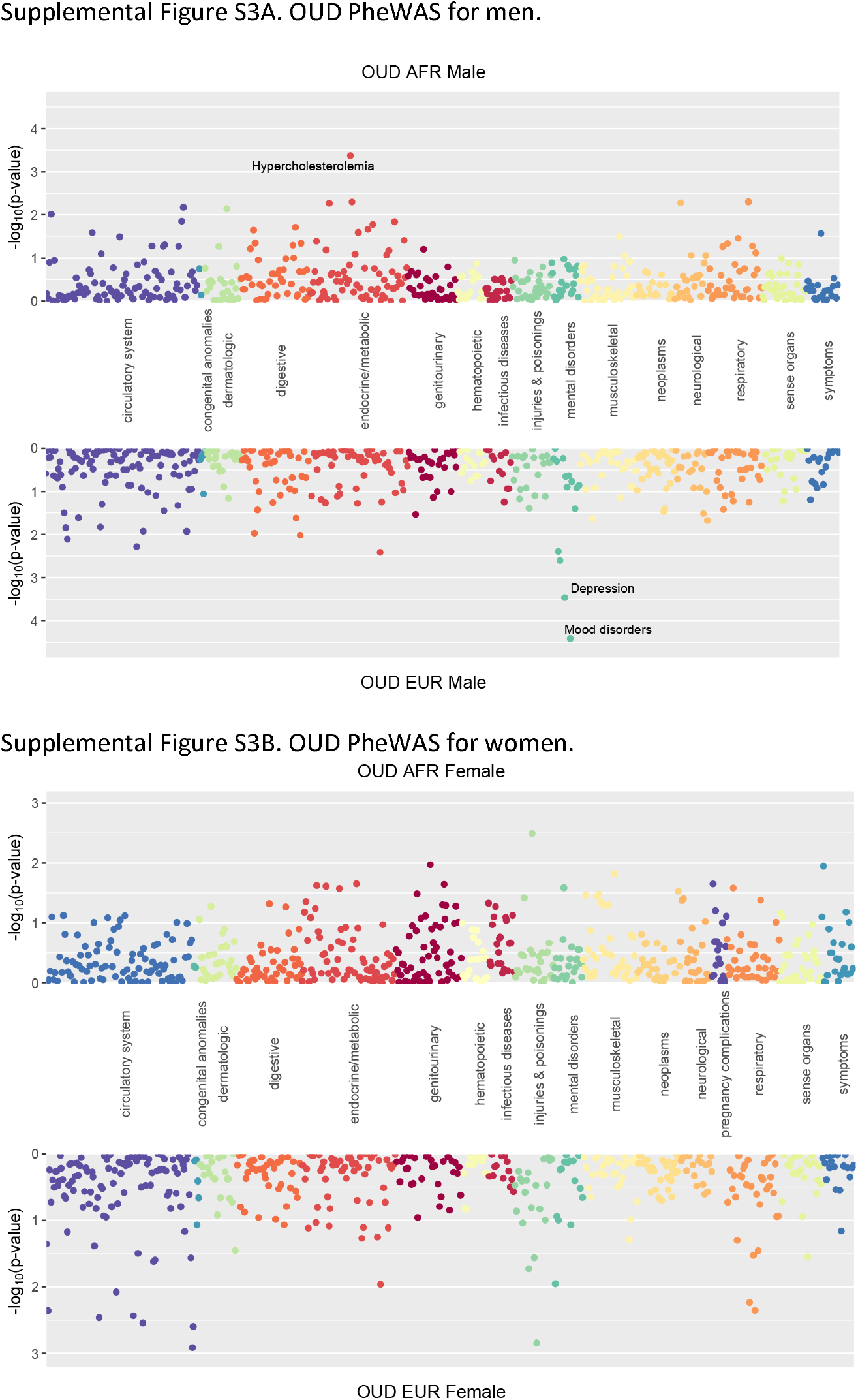
Phenome-wide association of polygenic risk scores for opioid use disorder (OUD) by sex for each ancestry group.

